# A Machine Learning-enabled SIR Model for Adaptive and Dynamic Forecasting of COVID-19

**DOI:** 10.1101/2024.07.30.24311170

**Authors:** Peter Mortensen, Katharina Lauer, Stefan Petrus Rautenbach, Marco Gallotta, Natasha Sharapova, Ioannis Takkides, Michael Wright, Mathew Linley

## Abstract

The COVID-19 pandemic has posed significant challenges to public health systems worldwide, necessitating accurate and adaptable forecasting models to manage and mitigate its impacts. This study presents a novel forecasting framework based on a Machine Learning-enabled Susceptible-Infected-Recovered (ML-SIR) model with time-varying parameters to predict COVID-19 dynamics across multiple geographies. The model incorporates emergent patterns from reported time-series data to estimate new hospitalisations, hospitalised patients, and new deaths. Our framework adapts to the evolving nature of the pandemic by dynamically adjusting the infection rate parameter over time and using a Fourier series to capture oscillating patterns in the data. This approach improves upon traditional SIR and forecasting models, which often fail to account for the complex and shifting dynamics of COVID-19 due to new variants, changing public health interventions, and varying levels of immunity. Validation of the model was conducted using historical data from the United States, Italy, the United Kingdom, Canada, and Japan. The model’s performance was evaluated based on the Mean Absolute Percentage Error (MAPE) and Absolute Percentage Error of Cumulative values (CAPE) for three-month forecast horizons. Results indicated that the model achieved an average MAPE of 32.5% for new hospitalisations, 34.4% for patients, and 34.8% for new deaths, for three-month forecasts. Notably, the model demonstrated superior accuracy compared to existing forecasting models with like-for-like disease metrics, countries and forecast horizons. The proposed ML-SIR model offers a robust and adaptable tool for forecasting COVID-19 dynamics, capable of adjusting to new time-series data and varying geographical contexts. This adaptability makes it suitable for localised hospital capacity planning, scenario modelling, and for application to other respiratory infectious diseases with similar transmission dynamics, such as influenza and RSV. By providing reliable forecasts, the model supports informed public health decision-making and resource allocation, enhancing preparedness and response efforts.

## Introduction

The COVID-19 pandemic has resulted in widespread economic, health and societal implications. To date, more than 700 million COVID-19 infections have occurred resulting in over 7 million deaths (The World Health Organization, 2024). COVID-19 reached pandemic status in March 2020 (The World Health Orgization, 2020), prompting most countries to introduce strict containment measures to mitigate its spread as well as surveillance systems to monitor epidemic changes. Infection-derived immunity and the introduction of COVID-19 vaccinations have reduced the risk of severe infections and mortality and have led to a withdrawal of the status ‘epidemic of public health concern’ by the World Health Organisation (WHO) in May 2023. Consequently, national surveillance systems have been reduced with notable changes in testing and reporting rates making it difficult to monitor disease dynamics over time.

Despite these advancements, SARS-CoV-2 has continued to cause periodic waves of increased COVID-19 cases. The dynamics of the pandemic have continually shifted due to the emergence of new variants, the deployment of new vaccines and treatments, and evolving policy responses. Periods of increased infection rates have resulted in higher rates of hospitalisation and deaths, which continue to pose significant threats to public health. Forecasting future cases, hospitalisations, and deaths using established epidemiological methods has proven challenging and often inaccurate. Accurate forecasting is crucial for informing post-pandemic policies, the production and stockpiling of prophylactics and treatments, and enabling first-line health responders to assess patient needs and resource demands effectively.

Current COVID-19 forecasting models have faced numerous challenges. Traditional models, such as those based on the susceptible-infected-recovered (SIR) framework, often fail to account for the complex and evolving nature of the pandemic. For instance, the model developed by Imperial College London, which was initially influential, faced criticism for overestimating hospitalisations and deaths due to its assumptions and parameter estimations (Flaxman et al., 2020). Similarly, models relying heavily on data from initial outbreaks struggled to adapt to new data reflecting changing virus transmissibility, virulence and public health interventions (Ioannidis et al., 2020). These models’ limitations underscore the need for more adaptive and robust forecasting tools.

The method proposed in this paper creates a framework to reliably forecast COVID-19 disease dynamics by efficiently adapting to the reported time-series data in each country, using an SIR model with time-varying parameters, with emergent patterns being forecasted forward, to estimate the new cases, new hospitalisations, hospitalised patients and new deaths.

The proposed method aims to address these challenges by creating a framework capable of reliably forecasting COVID-19 disease dynamics across multiple geographies, where data are reported. This is achieved by efficiently adapting to the reported time-series data in each country using a novel machine learning-enabled SIR model with time-varying parameters. The model forecasts emergent patterns to estimate new cases, new hospitalisations, COVID-19 patients in hospital, and new deaths. By incorporating machine learning techniques, the model can dynamically adjust to new data, improving accuracy and reliability in predicting future COVID-19 trends.

## Methods

### Data

Time-series data from 01/01/2020 to 20/02/2024 reported for COVID-19 new cases, new hospitalisations, hospitalised patients (the number of patients in hospital at the reporting date), and new deaths were collected for five countries. Data for the United Kingdom, Italy, Japan, and Canada, available under a CC-BY-4.0 license, were sourced from Our World in Data (OWID) as of 20/02/2024 (Edouard Mathieu, 2020). The data for the USA, made available under a Public Domain U.S. Government license, were collected from the CDC for all metrics (Centres for Disease Control and Prevention, 2024). All data obtained from the different sources were ingested into an internal database, structured and standardised. This collected data then goes through a preprocessing standardisation process. Preprocessing involved scaling the data to reflect daily data and interpolating the data onto daily time points. Then data was further smoothed with a 7-day rolling average.

### Compartmental Model

Compartmental models are essential tools in the study of the dynamics of dependent sub-populations, from ecology to epidemiology. In the specific case of epidemiology, they are commonly referred to as SIR models, after the most fundamental form with three sub-populations; susceptible, infected, recovered. These models help in understanding the spread and control of diseases within a population.

In our proposed method the compartmental model includes five key sub-populations, namely: the susceptible group, *S*, the exposed (or incubating) group, *E*, the infectious group, *I*, the hospitalised group, *H*, the recovered group, *R*, and the deceased group, *D*. Also, three non-conventional subpopulations, *I*_*2*_, *H*_*2*_, and *L* have been included to find the cumulative infections, hospitalisations and discharges from hospital (or leaving hospital), which can in turn be used to find the daily new cases and daily new hospitalisations and daily hospital discharges. *I*_*2*_, *H*_*2*_ and *L* do not appear in the right-hand side of the system of equations (i.e. they do not inform the predicted results). The various flow rates between these groups are illustrated in the schematic below in Fig. 1. With the model ordinary differential equations (ODEs) being defined as:

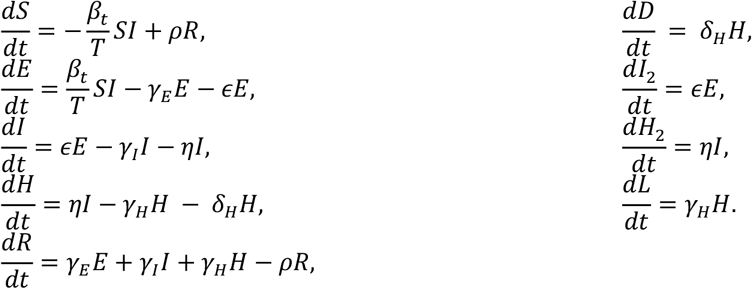

**Figure 1.**
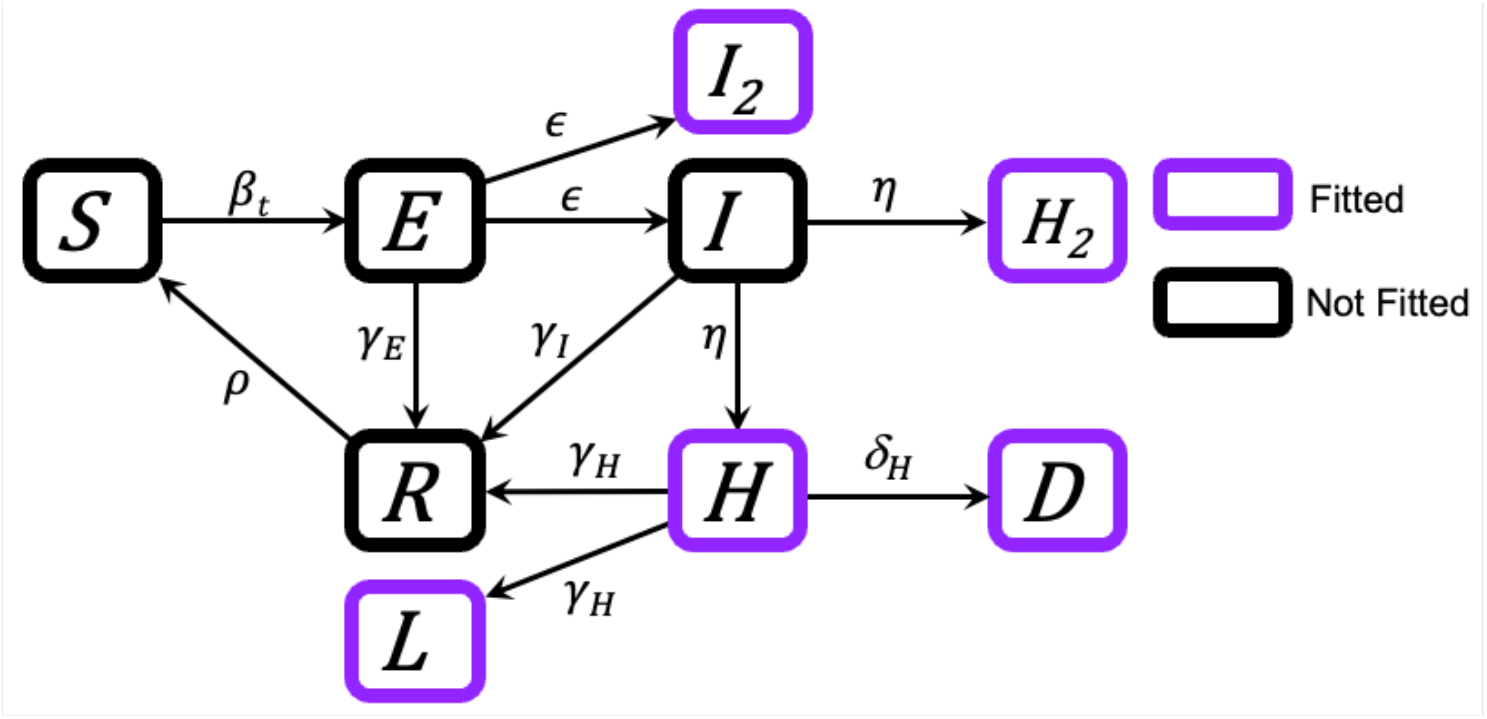
Schematic of the compartmental model, with purple boxes denoting compartments with which equivalent reported data can be used to fit the model parameters.

**Figure 2.**
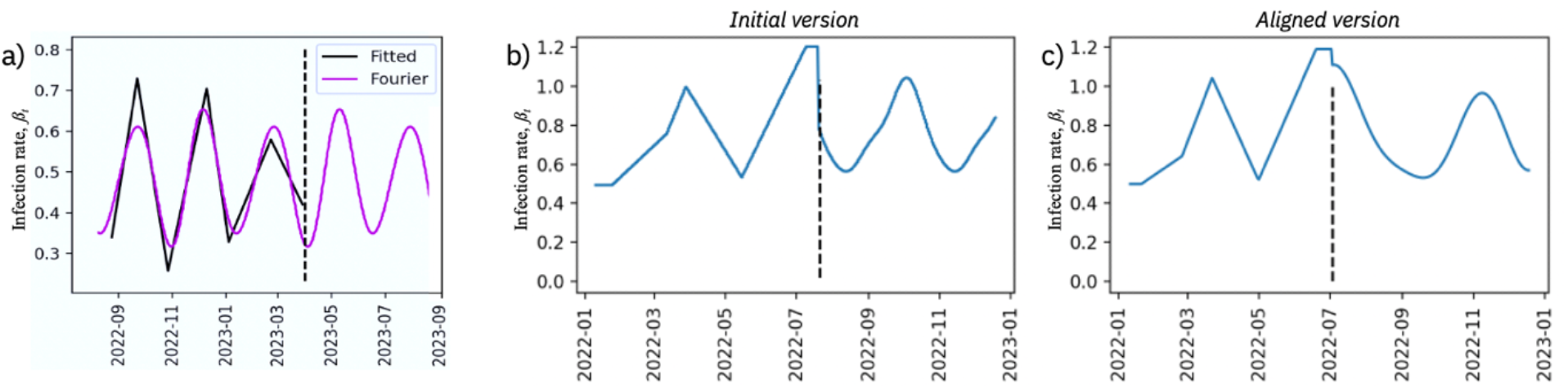
In each plot the vertical dashed line represents the end of the training data a) Infection rate parameter fitted from trends in reported data and third order Fourier series fitted to the fitted infection rate. The Fourier series was then forecasted beyond the observed training data. b) Initial version of transition point from fitted infection rate parameter to forecasted infection rate from Fourier series. c) Aligned version of transition point from fitted infection rate parameter to forecasted infection rate from Fourier series.

The above parameters of the ODEs are defined as: the time-varying infection rate, *βt* (where *t* is the time step), the rate of becoming infectious, 0, the rates of recovering of the exposed group, the infectious group, and the hospitalised group, *γ*_*E*_, *γ*_*I*_, and *γ*_*H*_ respectively, the rate of hospitalisation, *η*, the rate of becoming susceptible again, *ρ*, and the rate of death. It is difficult to measure these parameters as they have rate dimensions “per-time”, and thus the values of these parameters have been found through a parameter fitting algorithm by fitting the model to existing data, specifically new cases, new hospitalisations, new deaths and hospitalised patients. These metrics are equivalent to the compartments I_2_ H_2_ H and D, respectively. The parameter fitting process is described further in the below section “Parameter Fitting”. It should be noted that not all of the reported data are required to complete the parameter, although the parameter estimations are more robust where more disease metrics can be included. In the instances where new hospitalisations, patients and new deaths were available, hospital discharges were then calculated and used for further parameter fitting.

### Parameter Fitting

To estimate the parameters of the compartmental model, an established parameter fitting algorithm was used, specifically, the least-squares (SciPy, 2024). This function was chosen as it can robustly fit non-linear curves to data using the Trust Region Reflective algorithm which can handle large sparse problems with bounds on the parameters. This is done by finding a local minimum of a cost function provided for the relevant parameter set. In this instance the function finds the residual between select model outputs and the respective reported data. The residual can be built using new cases, new hospital admissions, new deaths and hospitalised COVID-19 patients. Commonly, not all the metrics are reported, however it is not necessary for all the data points to be used, and also since new cases are reported inconsistently as rates of testing and reporting are decreasing it has been found that fitting and forecasting is best improved by omitting the metric from the residual when there is sufficient data from the other metrics. In the instances in which, new hospital admissions, patients and new deaths are available, a new metric *discharges* can be estimated by assuming the changes in the number of patients is equal to the difference between the new hospitalisations and the new deaths and discharges. Thus,

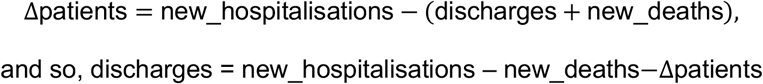

Although this metric is neither reported nor used in further analysis, it is beneficial to the parameter fitting process, as it more robustly defines the dynamics of the disease progression. As an example in a poorly defined system, an increase in say the hospitalisation rate and an increase in the hospital recovery rate will result in the same net number of patients, however this is mitigated through more complete data sets. Additionally, the model parameters are limited through the enforcement of reasonable bounds, based on their expected magnitude and thus further strengthening the parameter fitting.

### Time-varying Parameters

As previously discussed, the disease dynamics of COVID-19 are constantly changing due to changing pathogen characteristics such as new variants with different infectiousness and virulence, changes in herd immunity and other external factors. Therefore, to cover a time-period sufficient to train the model, time-varying parameters were used. Specifically, the infection rate, *β*_*t*_, was allowed to vary over time. Through experimentation it was found that the optimal time between peaks is approximately 40 days for training purposes, which was determined through trial and error.

The length of the training period was determined by the time between parameter changes, ensuring an even frequency for the time-vary parameter. In the validation process the training period was 160 days (a factor of 40). With the infection rate, *β*_*t*_, defined at six distinct time points,

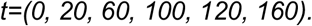

The first and final time points were fixed, but, for optimal parameter fitting, the intermediate time points were allowed to vary and fitted in the same parameter fitting process described previously. This required a minimum and maximum bounds for these time points, along with their initial values. These were defined to be,

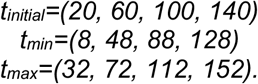

This allows the times at which the parameters vary to be adjusted, without the individual time points to touch or overlap, i.e. the second time point was initially at t=20 and bound between t=8 and t=32, the third time point had a minimum bound of t=48. The values of the minimum and maximum of the time points were 30% above and below the initial value (relative to the time between the points). For example, for the initial time point at 20 days, the maximum bound was 20 + (0.3 × 40) = 32.

Using this set up for time-varying infection rate (with the other parameters remaining static) a greater fit was achieved on the training data, than using entirely static parameters. The resulting time-varying parameters created an oscillating pattern. A pattern that must be forecast forward, to effectively forecast the overall disease dynamics.

### Forecasting

To perform forecasts using the fitted compartmental model, the sample paths were run from the beginning of the training period, through the to the end of the 160-day training period, then were forecasted for an additional three months. The model uses both, the found static parameters and the found time-varying parameters. However, the time-varying parameters were not informed for the forecasted period, thus the found pattern of the infection rate must be forecasted first. Here, it was assumed that the following three months will follow the same pattern as in the training period, namely an oscillating pattern.

To capture the oscillating pattern, a Fourier series was used to estimate the behaviour of the infection rate, *β*_*t*_. Here the same parameter fitting process was applied to the parameters of a third order Fourier series, training on the fitted infection rates. Then the found Fourier series was used to project the pattern forward.

The Fourier series was defined as:

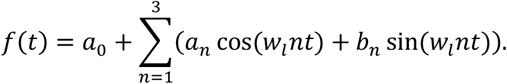

To make the transition from the found values, and the forecasted Fourier series values as smooth as possible, the Fourier series was shifted to minimise the difference at the points of transition.

An additional constraint was put upon the infection rate, *β*_*t*_, curve to ensure that the fitted Fourier series did not diverge out of the bounds of the original infection rates, *β*_*t*_, found during the parameter fitting stage in the training period.

Thus, if the Fourier series in the forecasted time-period was too large or too small the attempt was re attempted to find a more suitable estimate. Also, note that the infection rate must always be positive. Figure 3 shows an example of 50 runs in which several runs have extreme values for *βt* in the forecasted period, where the model is trained on data in the United Kingdom with the training period from the beginning of February 2023 to October 2023. The parameter fitting procedure found a number of Fourier series that were outside the boundaries of the originally fitted parameter values, thus they were excluded from the forecast.

**Figure 3.**
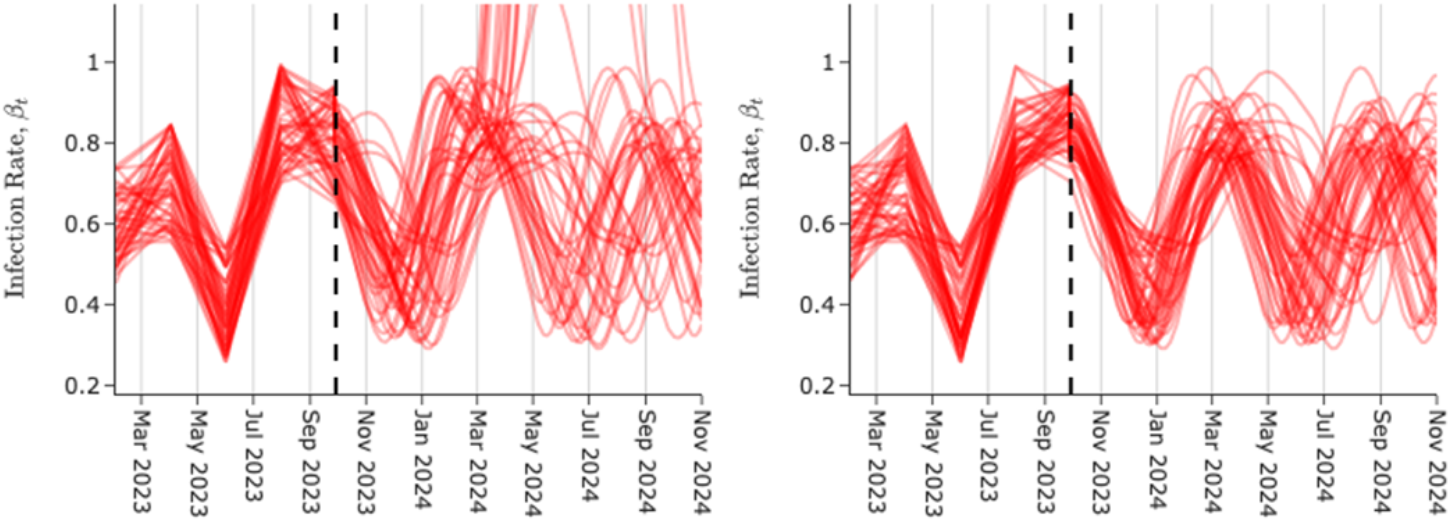
Examples of forecasted infection rate profiles, before (left) and after (right) having extreme results filtered out of potential runs.

Finally, it is important to note that the waves seen in the infection rate pre-empt the expected waves in the number of infections. This means that the infection rates towards the end of the training period were poorly defined, as they were informed by the waves outside of the training period. Thus, the final point of the found infection rate values was replaced by the mid-point value between the final and penultimate value, so that the final value did not distort the resultant Fourier series. Although the final point was included when the found infection rates were joined with the forecasted Fourier series, to ensure the final points of the training period aligned well with the training data, prior to forecasting.

The final forecast was constructed from 50 runs of the model, each resulting in a separate scenario, thus the median of these runs was used as the forecast, and the associated 50%, 80% and 95% prediction intervals were calculated.

### Assumptions and Limitations

The parameters of the model are found through a parameter fitting process, which relies on reported data. However, reported data is often not reflective of true values, with more recently reported data generally being less accurate than ‘older’ data, as recent data are often retrospectively updated (Lauren J. Beesley, 2022). Reported new cases are hugely dependent on testing rates with many infected people either choosing not to test or not being symptomatic enough to consider testing. For new hospitalisations and deaths, even though they are generally well reported the definitions of the “COVID-19 related hospitalisations or deaths” will vary between data providers but might also change within one source over the course of time (Tom Jefferson, 2022). Another key consideration is the timeliness, completeness and consistency of data reporting due to systematic inefficiencies or public holidays (Hildah Tendo Nansikombi, 2023). Also, over the course of the pandemic, sources have actively changed and updated their definitions for the reported disease metrics used to fit the model, and thus it is important to know what the definitions in the reported time-period refer to, and if there have been any changes to the definitions. As witnessed during the COVID-19 pandemic, testing rates are likely to degrade over time, particularly when the official status of an outbreak changes or effective treatments and prophylactics are introduced which may having a significant impact on the data quality over a specified training period. Therefore, if degrading testing rates are observed, and if the other metrics are reported well-enough, ‘cases’ are often removed from the fitting process. Overall, the model does not use any estimations of true values or any attempted standardisation of metric definitions, thus the reported results are relative to the standards of the reported data used (e.g. reported deaths are fed into the model with true-to-source reported values without normalisation for different definitions).

Another limitation is that the basis of the forecast assumes that the trends of the forecast will be the same as seen in the training period. This means it does not consider potential impacts of cases, hospitalisations or deaths from emergent new variants, increased contact events (i.e. the Christmas period), seasonal effects (i.e. increased time inside in the winter), governmental policies (i.e. lockdown procedures, which have been largely unused in the time periods considered and are unlikely to be significantly reintroduced), or increased vaccination schemes. It should be noted that adjustments could be made to estimate the impact of some external events) This model does not consider the population birth rate or the all-cause mortality rate, as it is assumed that the impact over the time-period being considered is negligible. Thus, the sum of the ODEs is zero, and the total population, *T*, is constant.

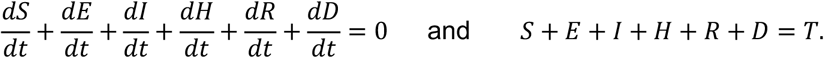

This model is referred to as a “late-stage” pandemic model as it is assumed that everyone who dies of COVID-19 passes away in hospital, which was not true in the early pandemic when a non-negligible number of people passed away outside of hospital, notably in care-homes.

Another key consideration in the construction of the compartmental model is there is no explicit value related to vaccinations. For COVID-19, vaccinations do not prevent infection or any later stage, thus a vaccinated person could not be isolated from potentially being infected and therefore should not form a separate compartment. However, vaccinations more broadly reduce the severity of outcomes, thus meaning vaccinations impact the magnitude of the fitted parameters. The model can naturally account for this due to the time-varying parameter fitting and a vaccine compartment was therefore excluded, however adding additional vaccination data into the model could potentially improve the control over the fitted parameters.

### Validation

The model was validated by back-testing the forecasts for the United States, the United Kingdom, Italy, Canada, and Japan; these countries were selected due to sufficient data availability. For each country 40 three-month forecasts were performed, a total of 200 forecasts. The first forecast from October 2022 and the final from February 2023, with a new forecast every three days between these two dates. This specific date range was chosen so that the training periods were always after the appearance of the omicron strains of SARS-CoV-2 and before the later stages of COVID-19 pandemic when the reporting of some metrics started to decrease. This meant the data used in the training period was the most consistent.

The set up of each of the 200 forecasts was the same, with the exception of which metrics were used for the training process (due to varying reporting standards from different countries). Each forecast comprised of 50 runs, each with unique, randomly selected (within the prescribed range) initial parameter estimations. For each run the parameters were fitted to the training data (the new cases data were not included as they were found to be too inconsistent) and the resultant fitted parameters were used to build a three-month forecast. The infection rate, *β*_*t*_, was allowed to vary in time with an initial choice of 40 days between parameter updates and the training period comprising of 160 days. To forecast the time-varying infection rate a Fourier series was fitted to the found *βt* parameter and projected forwards. With each of the 50 runs completed the associated median and 95%, 80%, and 50% prediction intervals were calculated. From the five countries that were used for the validation, the United States and Italy were completed using new hospitalisations, new deaths, and patients. The United Kingdom, Canada and Japan did not have new hospitalisation data. New cases were excluded from the fitting process, however the first time point in the training data was used to as the initial number of cases.

Validation metrics were calculated by comparing the forecast results with the observed data over the same respective time for each forecasting period. For each time point of the forecasted period the absolute percentage error between the observed value, *x*_*obs*_, and observed value, *x*_rep_, of each disease metric was calculated,

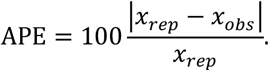

From this, two key error metrics were calculated, the mean absolute percentage error (MAPE) and the APE of the final time point of the cumulative metrics (CAPE). The MAPE is a measurement of how well the forecast fits to the reported data of the forecasted time-period, whereas the CAPE informs on the magnitude or the predicted cases, hospitalisations and deaths in the forecasted period. It should be noted that in the instances with no new hospitalisation data to validate against and patient data was used, the error calculation CAPE is not valid.

## Results

We visualised the retrospective forecast results, including sample paths and prediction intervals, along with the observed data for the United States, Italy, the United Kingdom, Canada and Japan for new hospitalisations, new deaths and patients (Figures 4, 5, 6, 7 and 8). In the forecasts for the United States and Italy for all three metrics the forecast results show good adherence to the training data and beyond into the forecasted period, following the trend observed in reported data with a magnitude close to the final points of data that were reported. For the United Kingdom, the forecasts for new deaths and patients follow the magnitude, timing and trends of the reported data well. It should be noted that, reporting of the new hospitalisation data in the UK ended in September 2022. Despite this the SIR model calculates a forecast for the new hospitalisations that has a magnitude consistent to the final reported values of the hospitalisations and the trends seen in the new deaths and patients. The resulting new hospitalisation forecast could not be validated against any data.

**Figure 4.**
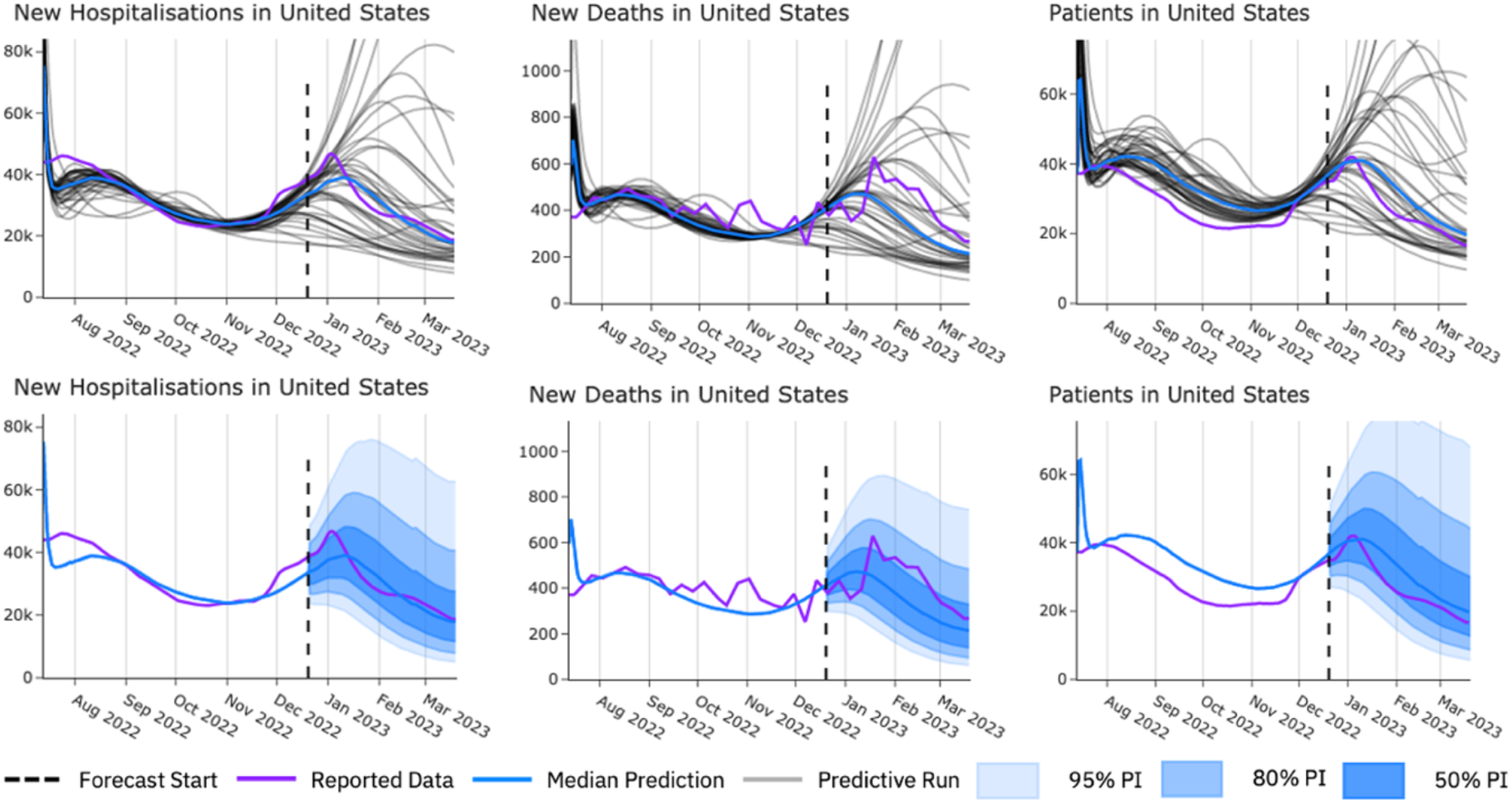
Example forecasts for United States for new hospitalisation, new deaths and patients. In each plot the vertical dashed line represents the end of the training data. Above, each predicted run’s sample path (grey), median prediction (blue) and observed data (purple). Below, median prediction (blue) and 95%, 80% and 50% prediction intervals (shaded blue) in the forecasted period compared with observed data (purple).

**Figure 5.**
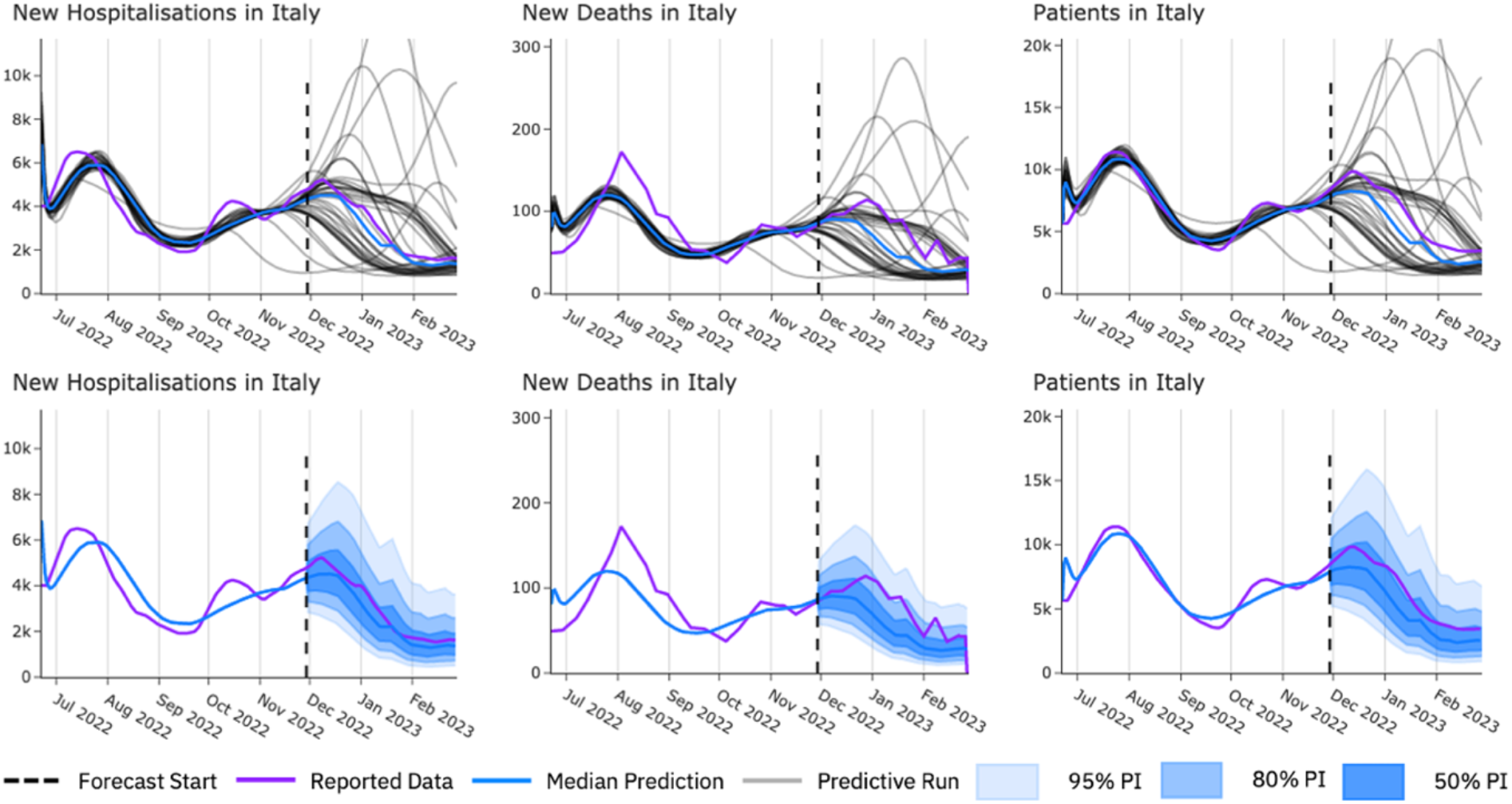
Example forecasts for Italy for new hospitalisation, new deaths and patients. In each plot the vertical dashed line represents the end of the training data. Above, each predicted run’s sample path (grey), median prediction (blue) and observed data (purple). Below, median prediction (blue) and 95%, 80% and 50% prediction intervals (shaded blue) in the forecasted period compared with observed data (purple).

**Figure 6.**
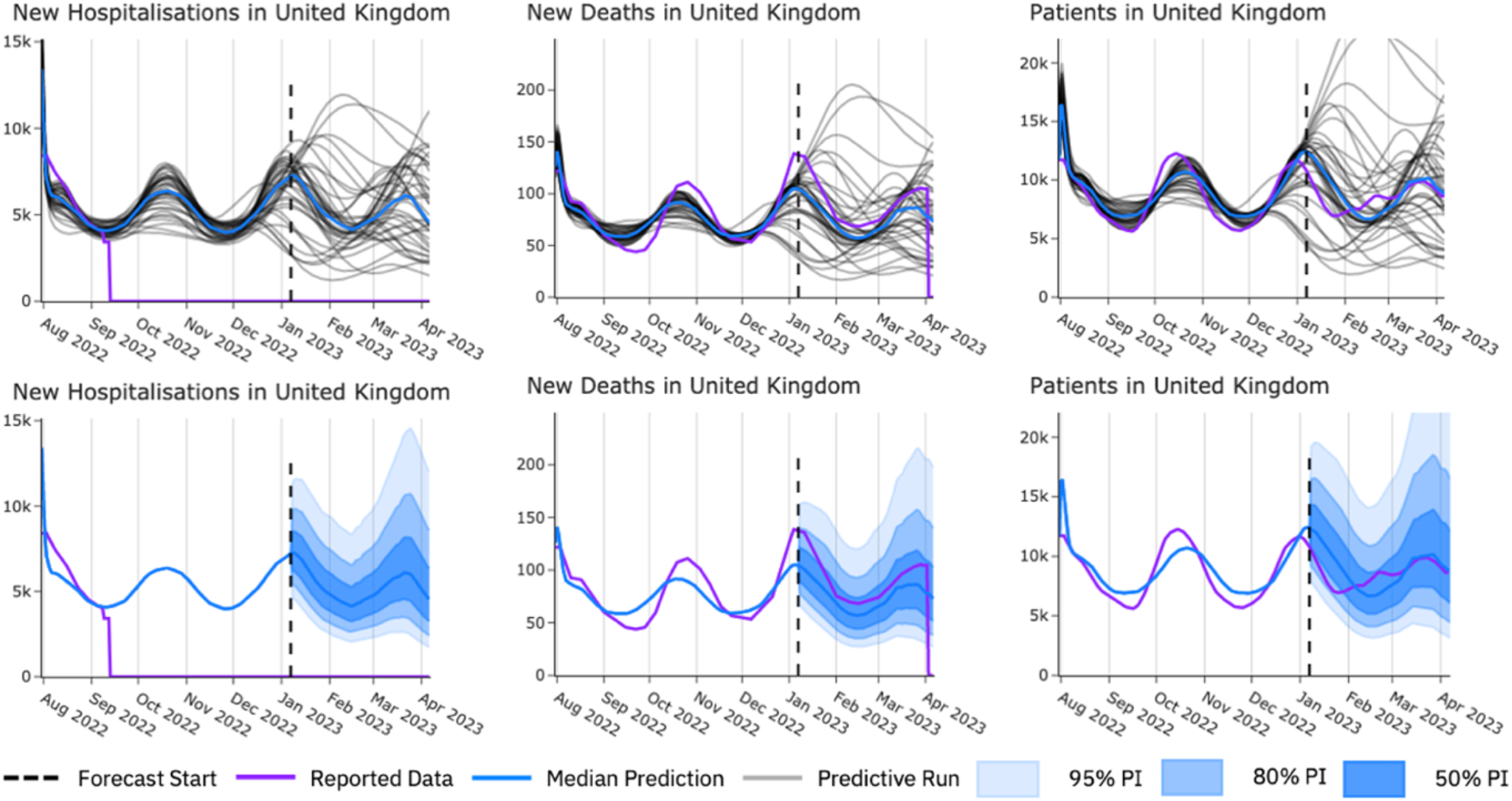
Example forecasts for United Kingdom for new hospitalisation, new deaths and patients. In each plot the vertical dashed line represents the end of the training data. Above, each predicted run’s sample path (grey), median prediction (blue) and observed data (purple). Below, median prediction (blue) and 95%, 80% and 50% prediction intervals (shaded blue) in the forecasted period compared with observed data (purple).

**Figure 7.**
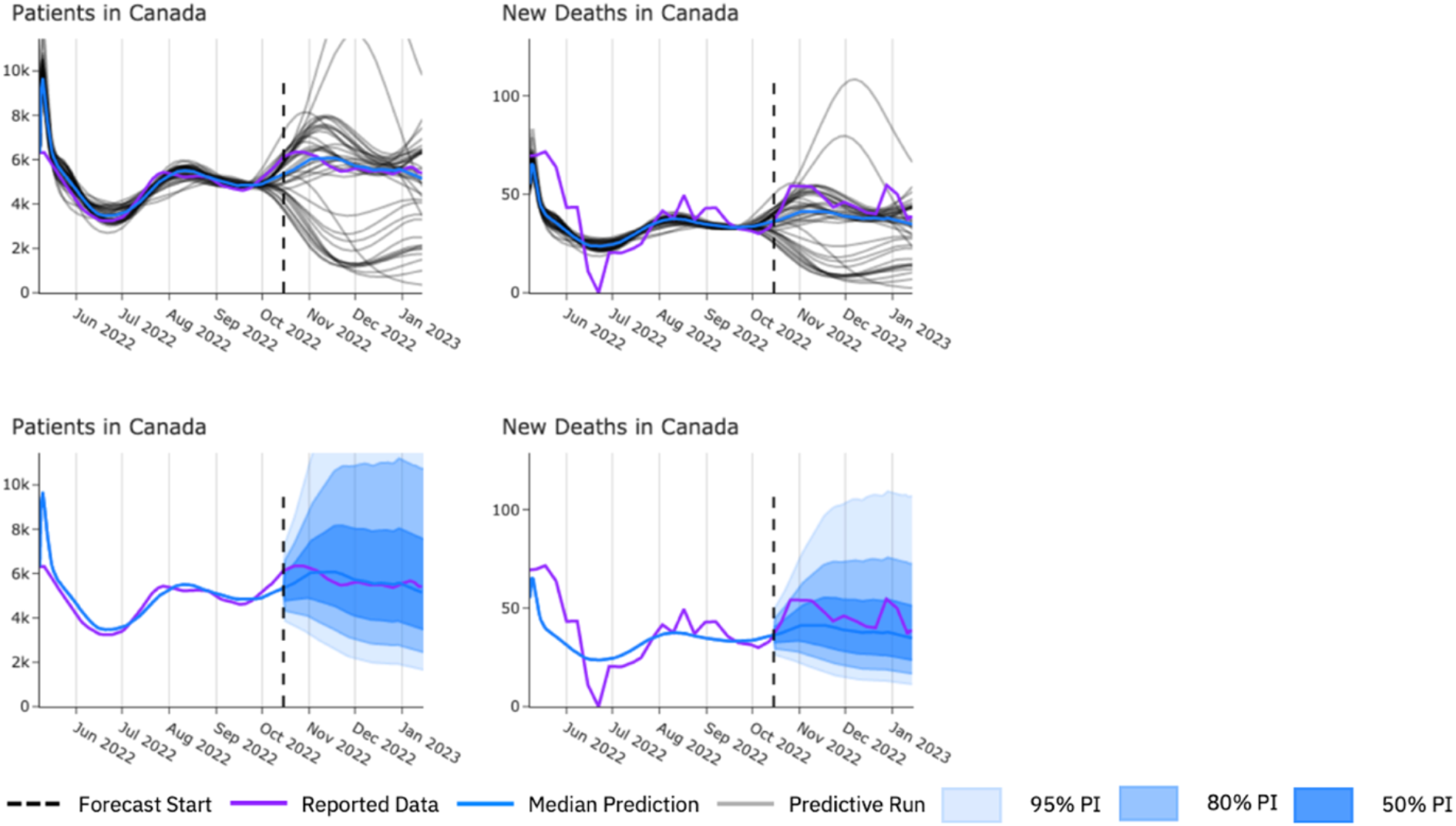
Example forecasts for Canada for patients and new deaths. In each plot the vertical dashed line represents the end of the training data. Above, each predicted run’s sample path (grey), median prediction (blue) and observed data (purple). Below, median prediction (blue) and 95%, 80% and 50% prediction intervals (shaded blue) in the forecasted period compared with observed data (purple).

**Figure 8.**
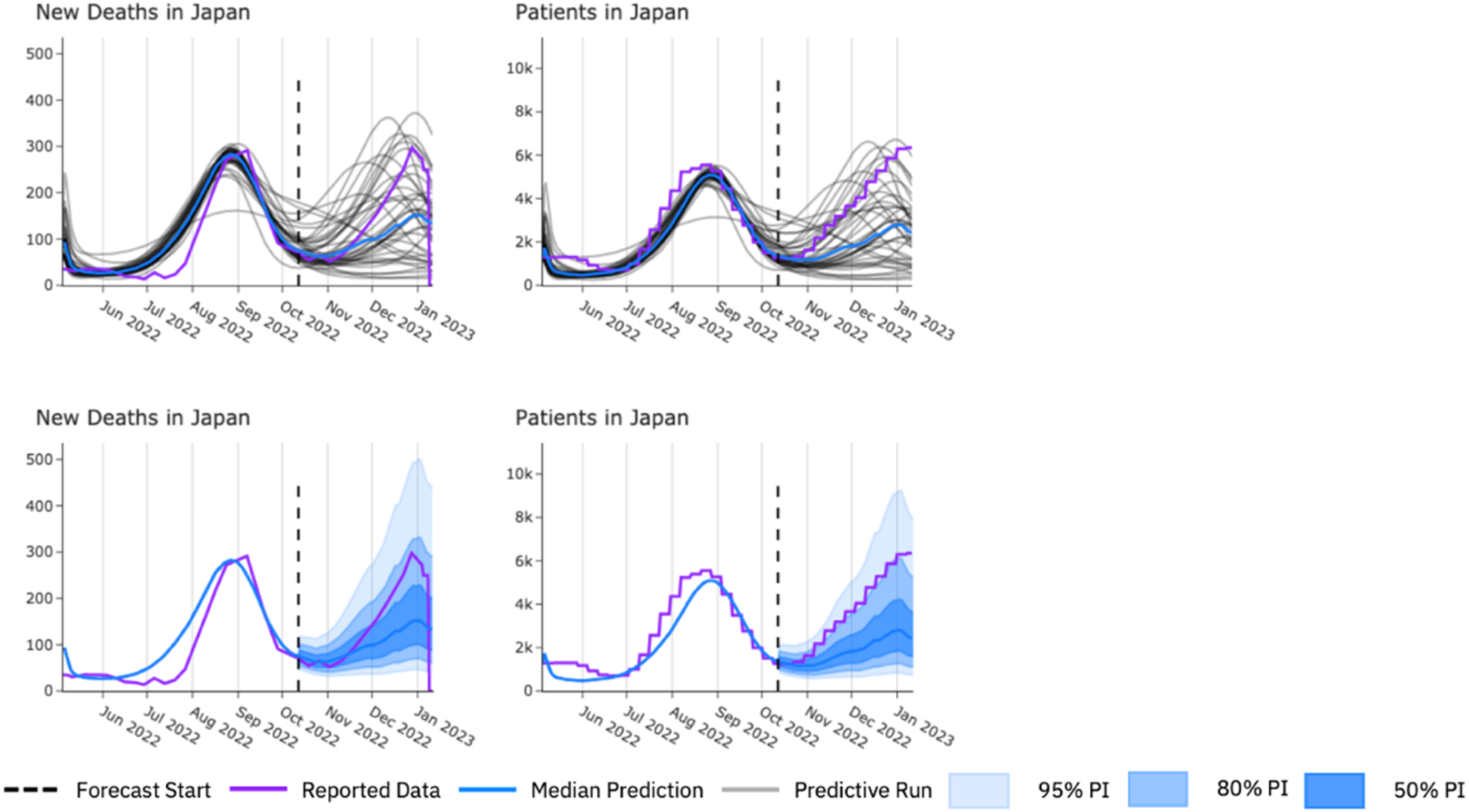
Example forecasts for Japan for new deaths and patients. In each plot the vertical dashed line represents the end of the training data. Above, each predicted run’s sample path (grey), median prediction (blue) and observed data (purple). Below, median prediction (blue) and 95%, 80% and 50% prediction intervals (shaded blue) in the forecasted period compared with observed data (purple).

The proposed forecasting framework was evaluated using data from the United States, Italy, the United Kingdom, Canada, and Japan. The accuracy of the forecasts was assessed using the Mean Absolute Percentage Error (MAPE) and the Cumulative Absolute Percentage Error (CAPE) across several metrics: new hospitalisations, patients, and new deaths (Table 1.). For new hospitalisations, only two of the countries had data available for validation. The MAPE for the United States was 20.9%, indicating a high level of accuracy. In contrast, Italy exhibited a higher MAPE of 32.4%. The overall MAPE for new hospitalisations across all countries was 26.7%. The CAPE for new hospitalisations was lower, with the United States at 19.7% and Italy at 26.0%, contributing to an overall CAPE of 22.9%.

**Table 1.** Validation results for three-month forecasts for each disease metrics for all countries and overall. Crosses mark where no data were available for validation.

|  | New hospitalisations |  | Patients |  | New deaths |  |
| --- | --- | --- | --- | --- | --- | --- |
|  | MAPE (%) | CAPE(%) | MAPE (%) | CAPE(%) | MAPE (%) | CAPE(%) |
| United States | 20.9 | 19.7 | 27.7 | X | 19.6 | 12.5 |
| Italy | 32.4 | 26.0 | 34.0 | X | 42.8 | 38.3 |
| United Kingdom | X | X | 43.0 | X | 39.8 | 32.1 |
| Canada | X | X | 33.3 | X | 35.2 | 30.5 |
| Japan | X | X | 33.8 | X | X | X |
| Overall | 26.7 | 22.9 | 34.4 | X | 34.4 | 28.4 |

The accuracy of patient forecasts was also evaluated. The MAPE for the United States was 27.7%, while Italy, Canada and Japan had a MAPE of between 33-34%. The overall MAPE for patient forecasts was 34.4%. The new deaths metric showed larger variation in forecast accuracy. The United States had a MAPE of 19.6% and a CAPE of 12.5%, demonstrating high accuracy. Italy, on the other hand, had a MAPE of 42.8% and a CAPE of 38.3%. The United Kingdom, Canada, and Japan had MAPEs of 43.0%, 33.3%, and 33.8%, respectively, potentially highlighting that differences in reporting standards for deaths may yield variable results. The overall MAPE for new deaths across all countries was 34.4%, with an overall CAPE of 28.4%.

The forecasts were performed for a three-month time period. Figure 9 overviews the average MAPEs and CAPEs of the new hospitalisations, patients, and new deaths across all countries for a range of forecast durations, from two weeks up to 12 weeks (Figure 9.) As expected, the further forward the model forecasts the less accurate it becomes. However, an increase in forecast horizon does not see an equally proportional increase in error. For example, increasing the forecast horizon from four weeks to 12 weeks (a 300% increase) sees the MAPE for new hospitalisations increase from 17% to 27% (a 58% relative increase). Considering the same, the MAPE for patients increases from 27% to 34% (a 27% relative increase), and the MAPE for deaths sees a growth from 21% to 34.4% (a 63% relative increase). This highlights the stable performance of the model over longer forecasting horizons.

**Figure 9.**
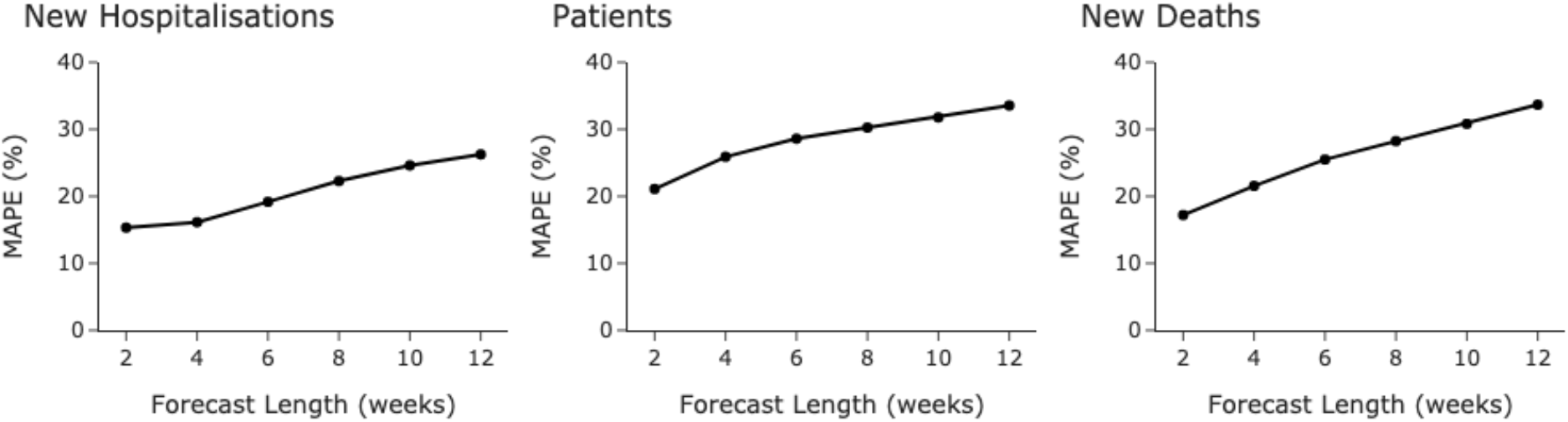
Overall mean absolute percentage error values for different forecast horizon lengths, from two to 12 weeks, across all five countries for each disease metric.

The MAPE values for the forecasts were compared against previously published COVID-19 forecasting models for the United States (Figure 10.). These are models that were submitted to the CDC for forecasting COVID-19 in the US, where the CDC then generated an ensemble model from the submitted models, which outperformed every other model in their study (Cramer, 2021). When directly comparing our US model performance with the CDC model, the error values are lower at all timepoints, including at three month forecast horizon compared with their four-week forecast.

**Figure 10.**
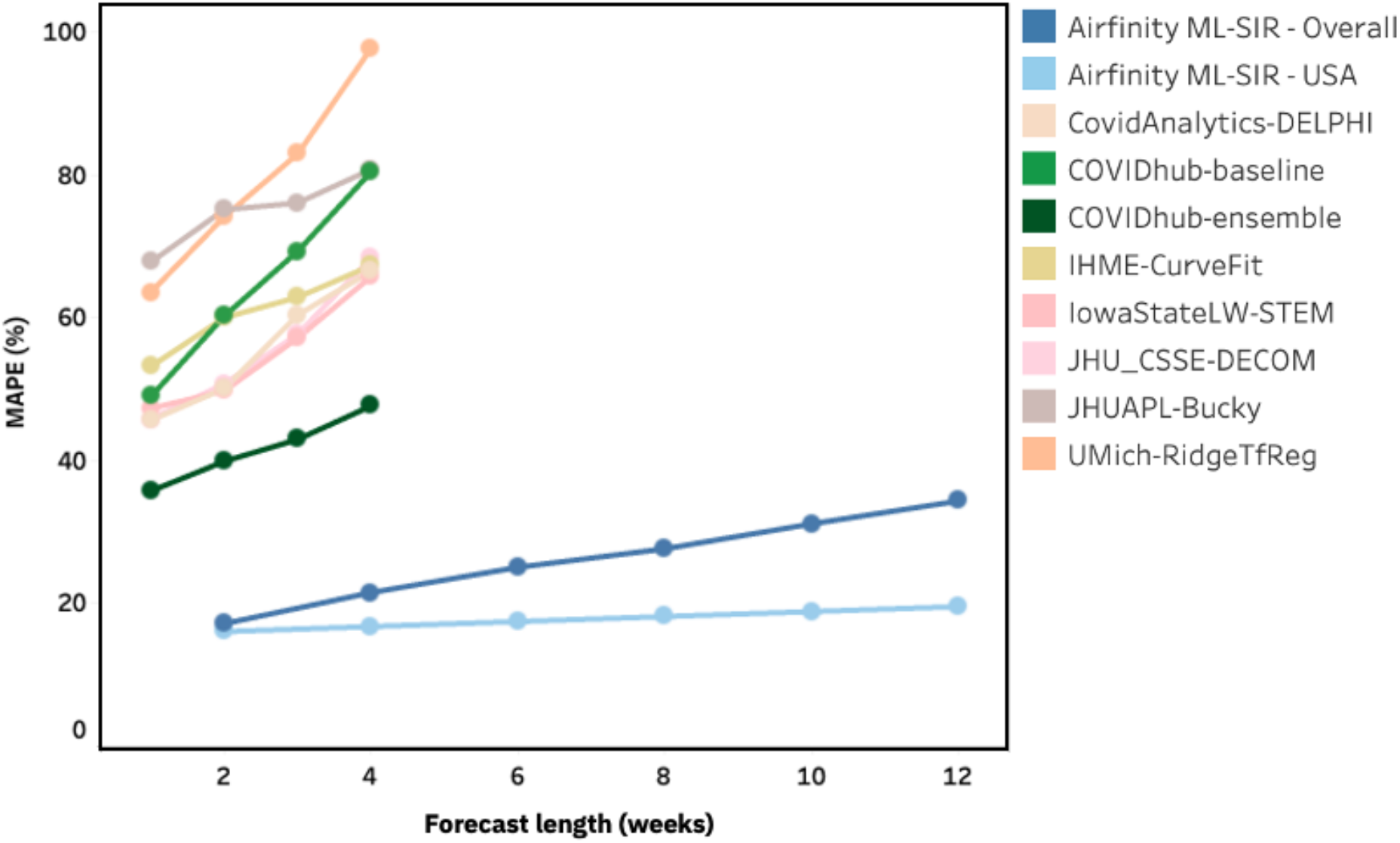
Comparison of MAPE values for deaths for different forecast horizon lengths (in weeks) for different models. Airfinity ML-SIR model results, showing the overall scores across the 5 countries and the scores for the USA specific forecasts, are compared with some of the models submitted to the CDC for US forecasts. Also shown is the CDC’s ensemble model from the submitted models, the COVIDHub-ensemble.

At a four-week forecast horizon for deaths the models submitted to the CDC study and the CDC ensemble model were compared with our model (Figure 11.). With a four-week forecast we achieved a MAPE of 16.8% for the USA compared to the 47.7% achieved by the CDC ensemble model. The average MAPE across all the submitted models was 80.6% (including models that had a MAPE of greater than 100%, which are not included in Figure 11.)

**Figure 11.**
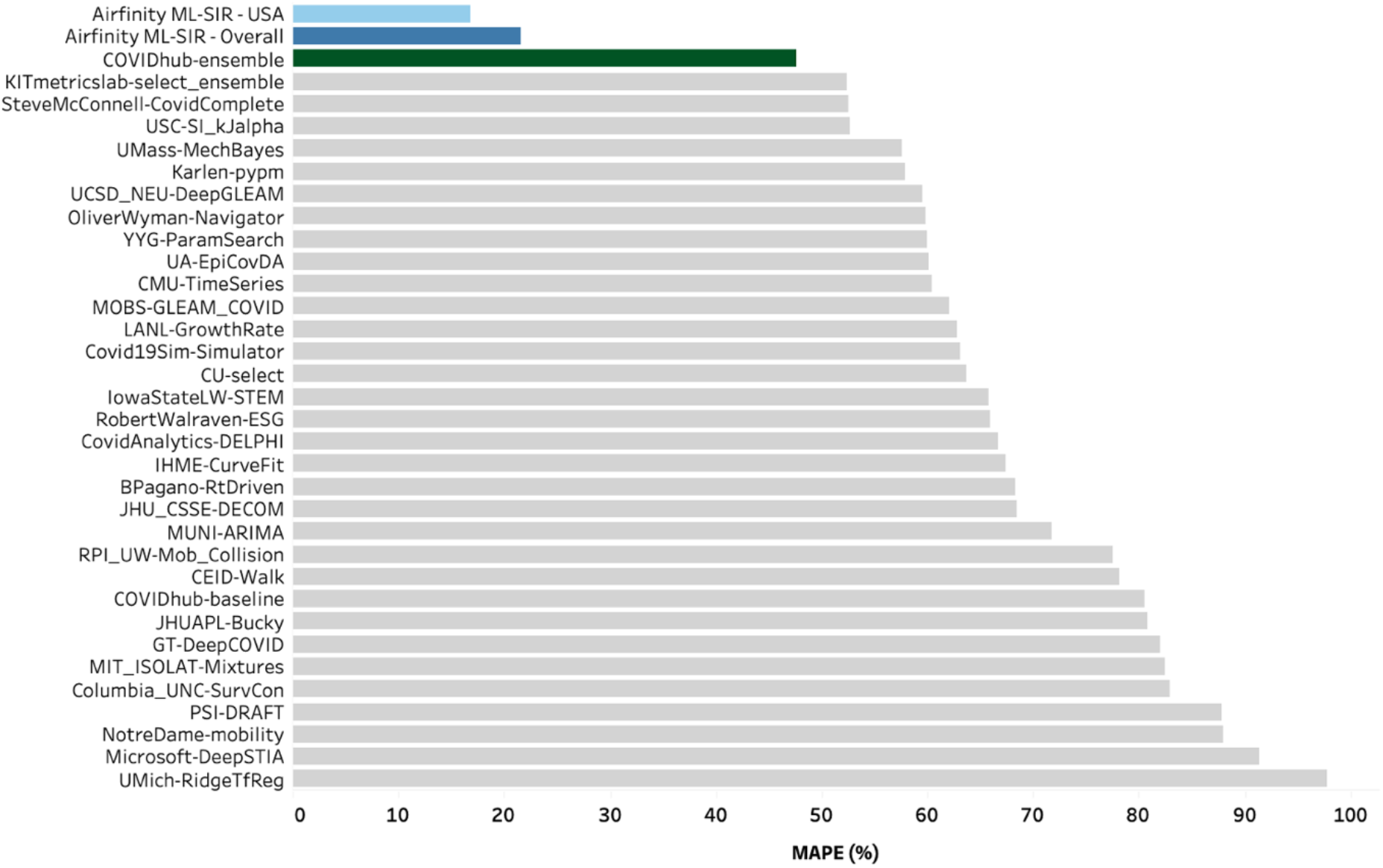
Comparison of MAPE values for deaths at four-weeks forecast horizon for different models. Airfinity ML-SIR model results, showing the overall scores across the 5 countries and the scores for the USA specific forecasts, are compared with the models submitted to the CDC for US forecasts (models with a MAPE of over 100% have been excluded from the visualisation). Also shown is the CDC’s ensemble model from the submitted models, the COVIDHub-ensemble.

## Discussion

Public health data on infectious diseases often suffer from quality and representativeness issues due to inconsistent surveillance, a lack of standardisation, timeliness, and resource limitations (Cristina Costa-Santos, 2021) (Groseclose, 2024). These aspects are further compounded by sociopolitical factors and privacy concerns, which can hinder accurate data collection and reporting, leading to challenges in the overall surveillance and response to infectious diseases (Ensheng Dong, 2020). Managing healthcare and protecting populations from crises such as the COVID-19 pandemic based on these data is difficult and requires flexible and adaptable models to provide forecasts that support decision makers.

The proposed framework in this study creates a highly versatile, dynamic forecast based on a range of metrics and countries, quickly adaptable to other scenarios and reporting standards. The resulting forecasts have been validated for the USA, United Kingdom, Japan, Canada, and Italy due to the best data availability. The novel framework details a method to fit the parameters of a late-stage COVID-19 SIR model, incorporating both static and time-varying parameters to account for the shifting dynamics of COVID-19 spread for any country with sufficiently reported data. A country’s data reporting is considered sufficient if at least two of the following metrics are reported: new cases, new hospitalisations, new deaths, or patients.

The validation process showed that the average mean absolute percentage error (MAPE) of forecasted new hospitalisations was 32.5%, and the average MAPE of forecasted new deaths was found to be 34.8%, both over a three-month forecast horizon. It should be noted that the forecasting framework consistently (and expectedly) overestimates new cases relative to the reported cases. This is likely due to the consistent decrease in testing. These estimations may form a basis for true case estimates; however, they cannot be validated without a better understanding of the true burden.

The performance metrics from our model highlight its adaptability, returning high accuracy across multiple geographies for longer forecast horizons. Individual forecasting models often show significant variability in accuracy, with performance fluctuating (John P.A. Ioannidis, 2022). An ensemble model coordinated by the US CDC (considered a benchmark for USA COVID-19 forecasting) was among the top performers and outperformed individual models. The average performance across the 41 individual models had a MAPE of 79.8%, compared with the ensemble performance of 47.7% for a four-week forecast horizon predicting new deaths (Cramer et al., 2021). When benchmarking our model against these results, it performs better with a MAPE of 16.8% under similar conditions (four-week forecast for deaths in the US). Furthermore, another SIR model that incorporated machine learning algorithms, known as SIMLR, showed MAPE values of 33% for four-week cases forecasts for the US, in line with the CDC COVIDHub-ensemble model (Roberto Vega, 2022). Notably, the authors of the SIMLR model note that their model performance was reliant on conditional probability values within the model that are hard to fit and thus were set manually using their domain knowledge, rather than a model fitting process. This further highlights the high degree of accuracy and adaptability of our ML-SIR model, without the need for manual parameter fitting.

As shown by (Cramer, 2021), models using additional data sources did not consistently show improved accuracy. These findings indicate that simply incorporating more data is not sufficient for better predictions, but instead requires any additional data to be incorporated in a considered manner. In our study we found that incorporating variables beyond the traditional SIR model led to an improvement of forecasts particularly when variation of parameters over time was allowed. A traditional SIR model implemented by Shams et al. (Fazila Shams, 2022) with a MAPE of 50.3% over a two-week forecast for Islamabad, while an SEIR model submitted to the CDC developed by UCLA had a four-week forecast MAPE of 133.4% for deaths in the USA (Cramer, 2021), compared with a MAPE of 16.8% for the US and 21.56% overall for the model we propose here. This is consistent with findings by (Vinay Kumar Reddy Chimmula, 2020), who demonstrated that integrating machine learning techniques with epidemiological models can enhance prediction accuracy by capturing the non-linear patterns in the data, however this model was highly specialised for forecasting within Canada alone.

It is well known from scientific literature that forecast accuracy and calibration degrade significantly with longer forecast periods. Often, the accuracy of the forecast drops beyond a four-week period. This phenomenon has been documented in multiple studies, including work by (Fotios Petropoulos, 2020), who highlighted the challenges in maintaining forecast accuracy over extended periods, especially in a rapidly evolving pandemic context. By capturing the trends in the non-linear time-varying parameters using Fourier series and forecasting those forwards, the prediction error for our forecasts remained low and even outperformed models with much shorter forecast horizons (Cramer 2021, Shams at al. 2022).

The proposed ML-SIR model’s adaptability has the potential to extend its utility beyond national forecasts to more localised applications, such as regional or hospital-level capacity planning. By incorporating local data, the model could provide targeted forecasts to help manage hospital resources, ensuring that facilities are prepared for surges in cases and hospitalisations. Additionally, the model can be adapted to incorporate granular age-specific data, enabling the analysis of the impact on particular risk groups, such as the elderly or those with comorbidities. This capability allows for more precise modelling of disease dynamics within vulnerable populations, informing tailored prevention and treatment strategies. Finally, the model could run different viral variant scenarios, adjusting the model parameters to reflect the transmissibility and severity of potential new strains. This feature allows health authorities to anticipate and prepare for the potential impacts of emerging variants, enhancing the resilience of public health systems against future disease surges.

Finally, we propose continued collaboration between public health agencies, governments, academic institutions, and industry partners to improve data quality and develop robust forecasting models to ultimately provide better healthcare for everyone. Further, there is a need to improve the predictive accuracy of models, especially by exploring adaptable and time varying models, as well as the integration of behavioural, mobility, and other real-time data streams, or relatively new data types such as wastewater data or crowd sourcing (Jihye Choi, 2016) (Rehab Meckawy, 2022) (Aminath Shausan, 2023).

## Conclusions

Here we proposed a novel and adaptable approach to forecasting COVID-19 disease dynamics. The primary benefit of the proposed ML-SIR model is its adaptability to new time series data and different geographies, while continuing to remain accurate over extended forecasting horizons. By using an SIR model with machine learning algorithms to determine time-varying parameters, the framework efficiently adapts to the reported time-series data in each country, allowing it to capture emergent patterns and forecast them forward. This adaptability ensures that the model can be applied to different countries, provided that sufficient data is available. Additionally, due to the machine learning component and the model’s adaptability, it is proposed that this model framework could relatively easily be applied to other use-cases as well as respiratory infectious diseases with similar transmission dynamics such as influenza and RSV. The model’s ability to adjust dynamically to new data inputs enhances its accuracy and reliability, making it a valuable tool for public health planning and response.

## Data Availability

All data produced in the present study are available upon reasonable request to the authors

## Notes

### Competing Interest Statement

All authors are current or previous employees of Airfinity Ltd.

### Funding Statement

This study did not receive any external funding

### Author Declarations

https://data.who.int/dashboards/covid19/

https://data.cdc.gov/Case-Surveillance/COVID-19-Case-Surveillance-Public-Use-Data-with-Ge/n8mc-b4w4/about_data

